# Proposal for a Value-Based Agreement to Reimburse Anti-obesity Medicines in Traditional Medicare

**DOI:** 10.1101/2025.07.10.25331289

**Authors:** Alison Sexton Ward, Erin Trish, Dana Goldman, Darius Lakdawalla

## Abstract

Anti-obesity medicines (AOMs), such as glucagon-like peptide-1 (GLP-1) agonists, hold great promise for slowing the chronic disease epidemic in the United States. Treatment with semaglutide and tirzepatide produces substantial weight loss and significant reductions in obesity-related diseases. Prior research shows expanding access to AOMs would generate trillions of dollars in social value to Americans today in the form of better life expectancy and fewer years spent with chronic diseases. The Medicare program, itself, could save around $245 billion in Part A and Part B spending over 10 years. Despite the potential value, access to GLP-1s for weight loss remains frustratingly low. Medicare and most *private* insurance plans do not cover the newly approved GLP-1 treatments. This paper presents a payment mechanism to resolve many insurers’ concerns regarding coverage of GLP-1s. We propose a value-based agreement for Medicare beneficiaries where The *Centers for Medicare & Medicaid Services* (CMS) pays a lower upfront price and shares the long-term savings with manufacturers if and when they are realized. For simplicity, we use a straightforward reimbursement approach building on Medicare’s existing risk-adjustment infrastructure to identify savings in treating obesity-related (ICD-10) conditions.

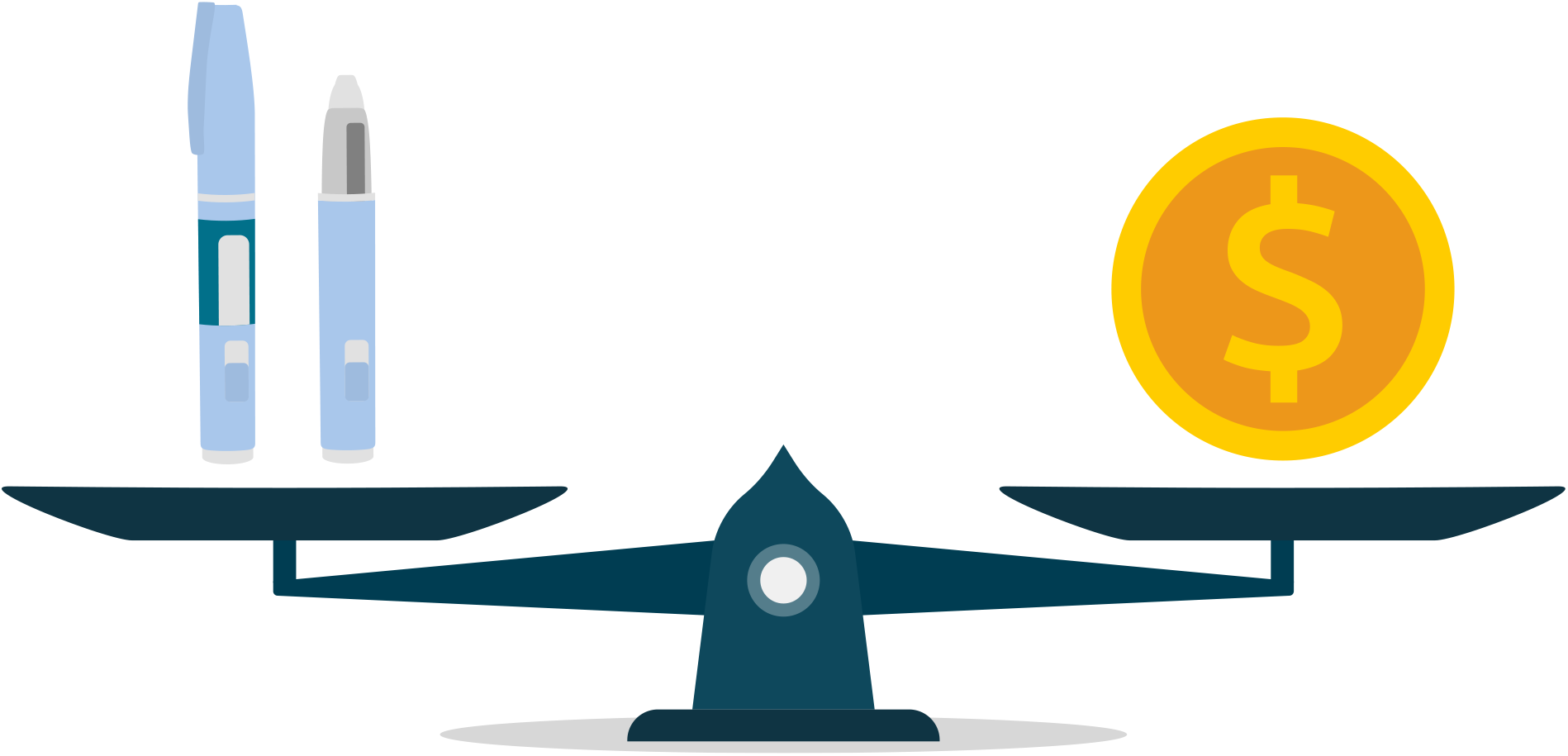

## Introduction

Anti-obesity medicines (AOMs), such as glucagon-like peptide-1 (GLP-1) agonists, hold great promise for slowing the chronic disease epidemic in the United States [1, 2]. The sequelae of obesity—including diabetes, heart disease, cancer and lung disease—affect 60% of U.S. adults, with 40% managing two or more conditions, and are responsible for 7 in 10 deaths each year [3, 4]. The financial toll is also staggering: Chronic disease accounts for approximately 90% of the nation’s $4.5 trillion in annual healthcare spending, largely driven by preventable hospitalizations, medication costs and long-term-care needs [4].

Fortunately for the 40% of Americans suffering from obesity, new AOMs offer hope. Treatment with semaglutide and tirzepatide produces substantial weight loss and significant reductions in obesity-related diseases [2, 5-7]. Elsewhere, we have found that expanded access to AOMs would generate trillions of dollars in social value to Americans today in the form of better life expectancy and fewer years spent with chronic diseases [8, 9]. Viewed as a public health investment, we find that expanded access to GLP-1s could provide annual rates of return on investment ranging from 13% to 22% [8].

The Medicare program, itself facing fiscal insolvency, could benefit, too: Our research suggests around $245 billion in offsetting Part A and Part B savings over 10 years [8]. Yet, despite the social return, access to GLP-1s for weight loss remains frustratingly low. Medicare and most private insurance plans do not cover the newly approved GLP-1 treatments. This paper presents a payment mechanism to resolve many insurers’ concerns regarding coverage of GLP-1s.

Payers’ reluctance to expand coverage for GLP-1s in part reflects concerns about long-term value.^1^ In particular, the industry has focused on real-world experience—especially adherence and discontinuation. Evidence suggests that up to 65% of weight-loss patients discontinue treatment in the first year and regain the lost weight [10, 11]. But skepticism is warranted. These analyses include older, less effective GLP-1s, and the results do not differentiate patients who discontinue for therapeutic reasons from those who discontinue due to lost insurance coverage, supply issues or a switch to cash pay. Nonetheless, the studies highlight the risk that insurers will pay for several months of expensive treatment without realizing sustained weight loss and future medical cost savings.

As highlighted by these discontinuation studies, payers and manufacturers disagree on the benefits. While clinical trial evidence for new GLP-1s like semaglutide and tirzepatide is clear— patients lost between 15% and 21% of their body weight on average [5, 6]—how that translates into a real-world, long-term benefit is uncertain. Payers and the patients they cover also suffer from asymmetry in their time horizons [12]. The benefits of weight loss extend well beyond most patients’ average tenure at an insurer, and this makes effective weight-loss therapies more valuable to patients than payers [13]. A grand challenge for policymakers is to overcome the narrow asymmetries of interests to obtain the socially desirable outcome: expanded access to GLP-1s.

We propose a value-based agreement (VBA) for Medicare beneficiaries using GLP-1s for weight loss. The Centers for Medicare & Medicaid Services (CMS) would enter into an agreement to pay a lower upfront price while sharing Medicare long-term savings with manufacturers if and when they are realized. These arrangements show promise in other clinical contexts, such as elevated cholesterol, where long-term treatment efficacy is uncertain [14-16]. But VBAs in the U.S. face challenges due to regulatory constraints, costly data collection and disagreements over financial terms [17]. Thus, we propose a straightforward reimbursement approach building on Medicare’s existing risk-adjustment infrastructure to identify savings in treating obesity-related (ICD-10) conditions. We outline how such a model would work below, but first we draw some lessons from Medicare’s prior experience with VBAs.

### Brief Lessons From Medicare’s History With Value-Based Care

In principle, VBAs offer two advantages: They provide a contractible solution when buyers and sellers disagree on the benefits of a health intervention, and they encourage marketplace actors to focus their efforts on improving patient outcomes instead of simply increasing the volume of goods or services provided [12]. In the GLP-1 weight-loss context, the first advantage addresses the uncertainty around long-term effectiveness described above. The second aligns Medicare and GLP-1 manufacturers around the goal of promoting appropriate use aimed at improving long-term health outcomes, addressing concerns about overuse for cosmetic weight loss [18].

To realize the promise of both advantages, a GLP-1 VBA must provide sufficiently powerful incentives to both sides. Tying too small a share of the payment to outcomes limits the VBA’s ability to bridge disagreements and limits marketplace incentives to curb overuse. Past experience with CMS VBAs from the 2010 Affordable Care Act provides a case in point. The Affordable Care Act mandated that CMS establish a pay-for-performance program for hospitals, which led to the Hospital Inpatient Value-Based Purchasing Program. This program sets aside a small portion of Medicare payments—2% as of 2025—as a bonus pool awarded to hospitals that outperform in those categories. In practice, the incentive payments have proven too small to have a significant impact on hospital care [19, 20].

In addition, a VBA must contract on outcomes that demonstrably matter to Medicare and its beneficiaries. The Merit-based Incentive Payment System (MIPS), established by the Medicare Access and CHIP Reauthorization Act (MACRA) of 2015, illustrates this point. In MIPS, clinicians earn a composite score across performance categories that adjusts their payments in a positive, neutral or negative way such that the program is ultimately budget neutral. However, a cross-sectional study of more than 80,000 primary care physicians participating in MIPS as of 2019 found scores to be inconsistently associated with performance on relevant process and outcome measures. In other words, MIPS was not incentivizing outcomes that really matter for quality of care [21].

Realizing the promise of VBAs for weight-loss treatment requires a substantial incentive component tied to salient outcomes. Tying outcomes-based payments to the avoidance of chronic disease presents a natural solution, because chronic disease imposes considerable burdens on patients themselves and on the Medicare program. In theory, tying financial penalties or rewards to diagnoses could influence the effort physicians spend diagnosing mild cases. However, this risk is mitigated in the traditional Medicare program because physician payments are reimbursed using services provided.

The traditional Medicare program is also well suited for an AOM VBA because CMS already measures the cost of chronic disease as part of its risk-adjustment methodology. CMS uses data from traditional Medicare to calculate the costs associated with various ICD-10 codes, which it uses to risk adjust payments in the Medicare Advantage program. While researchers debate whether this risk-adjustment methodology provides cost estimates relevant to Medicare Advantage [22], we propose using it here as an estimate of traditional Medicare costs associated with obesity-related diseases.

In sum, for both logistical and economic reasons, we propose locating the GLP-1 VBA program within traditional Medicare, contracting on the avoidance of chronic disease and measuring cost using Medicare’s existing risk-adjustment methodology. We estimate that expanding access to GLP-1s for weight loss could generate $175.6 billion to $245.1 billion in gross cost savings to Medicare over 10 years, enabling substantial, powerful incentive payments in the event of successful chronic disease prevention. Similarly, the prevalence of diabetes could fall by 5.5% to 9% over the same period.

### A Value-Based Contract for GLP-1 Therapies

We propose VBAs on GLP-1 therapies for weight loss. Based on current clinical criteria, this would provide AOM access to traditional Medicare beneficiaries with body mass index (BMI) of 30 or more, even without other GLP-1-qualifying chronic conditions. Conceptually, the monthly payment to the manufacturer would consist of three components: (1) a base payment per patient utilizing therapy in that month, (2) a copayment per patient utilizing therapy in that month and (3) a risk-based payment reflecting estimated Medicare savings accruing that month from prior use of GLP-1 weight-loss medicines. The base payment, copayment and share of savings paid to manufacturers would be negotiated between manufacturers and CMS. In keeping with the discussion above, we recommend that the at-risk portion comprise a substantial share of the potential compensation, and at-risk payments be linked to the avoidance of obesity-related chronic diseases. For simplicity of administration, we also recommend that monthly savings from each condition avoided be computed using Medicare’s existing risk-adjustment estimates of cost associated with specific hierarchical condition categories (HCCs), or a variant thereof. Figure 1 illustrates the arrangement. We focus on contracting for the product itself; additional considerations would be needed to ensure distribution (e.g., compensation to pharmacies and other entities in the distribution system or manufacturers including a proposed distribution model as part of their negotiations with CMS).

**Figure 1.**
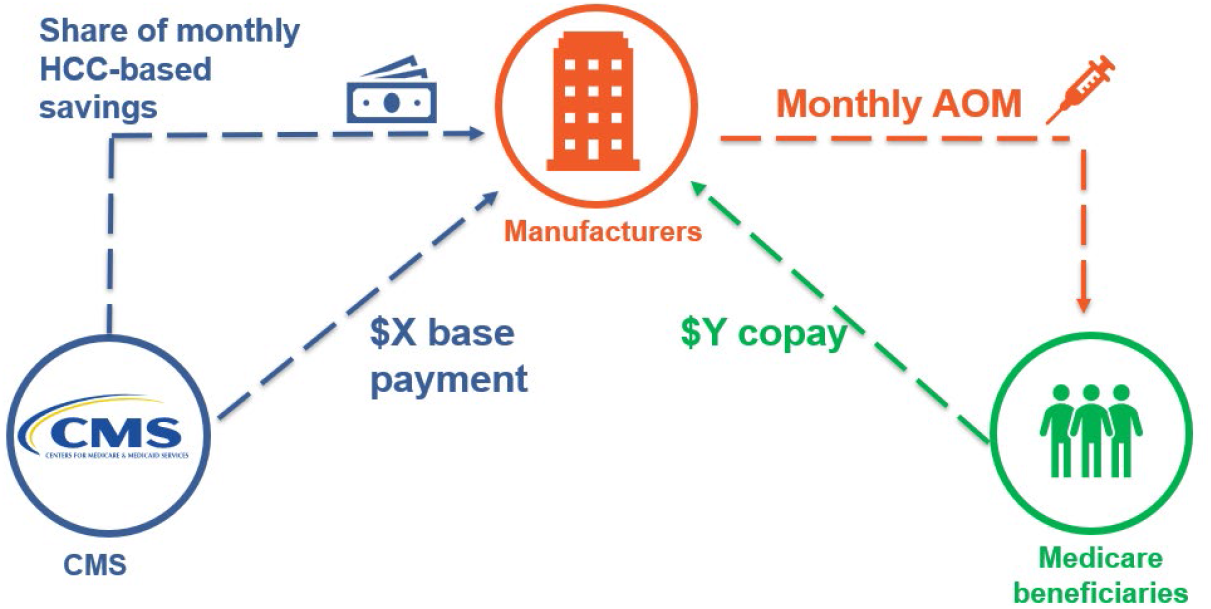
GLP-1 Value-Based Agreement Between CMS and Manufacturers

In the balance of this section, we discuss in further detail the specific contract parameters to be determined, the estimation of incremental savings from disease prevention, the estimation of disease cases avoided and an illustrative example of contracted payment terms.

### Contract Design

The negotiated VBA would need, at a minimum, to specify:

- the term, *T*, in months;
- the upfront payment, $X, that CMS will pay direct to manufacturers
- the monthly copay, $Y, that patients pay to manufacturers
- the set of “GLP-1-susceptible” conditions, *c* = {1, …, *C*}, drawn from the list of payment codes in CMS-HCC model;
- the set of demographic groups that CMS will use to stratify results, *d* = {1, …, *D*};
- the age groups covered in the agreement, *a* = {1, …, *A*};
- the method for computing the number of cases avoided due to GLP-1 usage;
- the share of the Medicare savings (potentially condition specific) that will be given to the manufacturer, *σ*_*c*_;
- and an agreement regarding the method for computing the incremental savings, *S*_*cccc*_, in month *t*, from avoiding a case of condition *c*, and for patients of age *a* in demographic group *d*.

### Incremental Savings

Manufacturers and CMS will need to agree upon a method for calculating the incremental savings, *S*_*ctda*_, across conditions, time, demographic groups and ages. A logistically simple approach leverages CMS’s existing HCC Risk Adjustment Model, which estimates traditional Medicare’s cost of treating specific HCCs. The HCC Risk-Adjustment Model aims to calculate the monthly capitated payments made to Medicare Advantage (Part C) plans for beneficiaries with different chronic conditions and demographic characteristics. The risk-adjustment model is based on a detailed mapping of ICD-10 codes to HCC codes. In its most recent version (V28), the CMS model maps 7,770 ICD-10 codes to 115 HCC codes that represent serious, costly or chronic conditions that impact payments. Many common or acute medical conditions like sprains or mild infections do not map to an HCC because they do not significantly affect costs over a year.

The CMS HCC codes are additive in nature and arranged into hierarchical groups such that CMS only counts the highest-severity code in its reimbursement (e.g., CMS would only risk adjust for heart failure rather than a less severe cardiovascular HCC if both were present). Each HCC is associated with a specific risk coefficient; however, the model also accounts for a few disease interactions that, when present together, have a significant impact on healthcare costs. The interactions are associated with their own coefficients that can be added to the individual condition’s coefficients. For example, type 2 diabetes and heart failure each have their own HCC coefficients, but there is also a coefficient for the disease interaction.

Finally, the CMS risk-adjustment model also adjusts for different patient demographics including age, sex, dual eligibility, disability and nursing home status. In fact, the HCC coefficients are stratified by nursing home status, disability and dual status such that the condition codes have separate coefficients for each of those statuses. To calculate a patient’s monthly capitated payment, the model adds all the HCC coefficients to get a total risk-adjustment factor score. An example patient risk-adjustment factor score is calculated in table 1.

**Table 1:** Example of Risk Adjustment Factor Score Calculations.

| Category | Description | Coefficient |
| --- | --- | --- |
| Demographic | Age 70, Female,<br>Community-Residing, Dual-Eligible Full Benefit | 0.506 |
| HCC Conditions | HCC37: Diabetes With Chronic Complications | 0.186 |
|  | HCC226: Heart Failure, Except End-Stage and Acute | 0.406 |
|  | HCC238: Specified Heart Arrhythmias | 0.407 |
|  | HCC280: Chronic Obstructive Pulmonary Disease (COPD) | 0.390 |
| HCC Interactions | Congestive Heart Failure + Diabetes Combination | 0.183 |
|  | Heart Failure*Chronic Lung Disorder | 0.109 |
| **Total Risk-Adjustment Factor Score** |  | **2.187** |

CMS uses the patient risk-adjustment factor score multiplied by county-level benchmarks to determine the maximum monthly payment available to Medicare Advantage plans. Benchmarks reflect both estimated traditional Medicare spending for an average-risk beneficiary in the county and a number of other adjustments reflecting Medicare Advantage payment policy. Here, we suggest using the county-level traditional Medicare spending estimates rather than incorporating the additional Medicare Advantage adjustments. Across all of the U.S. states in 2025, the monthly traditional Medicare spending estimates vary widely, from $776 in Presidio, Texas, to $2,433 in North Slope, Alaska, with a median value of $1,095 in Beaver, Pennsylvania.

The CMS-HCC model provides a ready-made structure on which to design a GLP-1 VBA, as it facilitates calculating the added Medicare costs of each obesity-related chronic condition. The V28 coefficient for diabetes ranges from 0.166 for community dwelling, non-dual beneficiaries who have aged into Medicare, to 0.28 for institutionalized (e.g., living in a nursing home) beneficiaries, which implies an added Medicare cost of $183 to $308 per beneficiary per month. Thus, if a base prevalence of diabetes among a certain Medicare population, say women ages 65–69, is determined, CMS can calculate the reduction in cases that occurred after expanding access to GLP-1s. The total Medicare savings from reducing diabetes could be estimated as the reduction in prevalence multiplied by the added Medicare cost (i.e., the HCC coefficient times the county-level traditional Medicare average spending). A similar calculation could be made for all obesity-related conditions to estimate the total Medicare savings. See table 2 for a list of obesity-related HCCs along with their coefficients for the different eligibility criteria (aged versus disabled), benefit types (non-dual versus dual and partial versus full benefit) and nursing home status (community dwelling or institutionalized).

**Table 2:** CMS HCC V28 Codes for Obesity-Related Conditions.

| Code |  | Description | Coefficients |  |  |  |  |  |  |
| --- | --- | --- | --- | --- | --- | --- | --- | --- | --- |
|  |  |  | CNA | CND | CFA | CFD | CPA | CPD | INS |
| Cancer |  |  |  |  |  |  |  |  |  |
| HCC 17 | Cancer Metastatic to Lung, Liver, Brain, and Other Organs; Acute Myeloid Leukemia Except Promyelocytic |  | 4.209 | 3.995 | 3.896 | 4.235 | 3.946 | 4.103 | 1.952 |
| HCC 18 | Cancer Metastatic to Bone, Other and Unspecified Metastatic Cancer; Acute Leukemia Except Myeloid |  | 2.341 | 2.486 | 2.277 | 2.537 | 2.166 | 2.403 | 1.11 |
| HCC 19 | Myelodysplastic Syndromes, Multiple Myeloma, and Other Cancers |  | 1.798 | 1.989 | 1.563 | 1.661 | 1.52 | 1.554 | 0.957 |
| HCC 20 | Lung and Other Severe Cancers |  | 1.136 | 0.978 | 1.166 | 1.173 | 1.214 | 1.067 | 0.672 |
| HCC 21 | Lymphoma and Other Cancers |  | 0.671 | 0.54 | 0.654 | 0.739 | 0.627 | 0.618 | 0.493 |
| HCC 22 | Bladder, Colorectal, and Other Cancers |  | 0.363 | 0.366 | 0.382 | 0.409 | 0.41 | 0.351 | 0.314 |
| HCC 23 | Prostate, Breast, and Other Cancers and Tumors |  | 0.186 | 0.233 | 0.196 | 0.218 | 0.203 | 0.237 | 0.197 |
| Diabetes |  |  |  |  |  |  |  |  |  |
| HCC 36 | Diabetes with severe acute complications |  | 0.166 | 0.191 | 0.186 | 0.235 | 0.166 | 0.21 | 0.28 |
| HCC 37 | Diabetes with chronic complications |  | 0.166 | 0.191 | 0.186 | 0.235 | 0.166 | 0.21 | 0.28 |
| HCC 38 | Diabetes with Glycemic, unspecified, or no complications |  | 0.166 | 0.191 | 0.186 | 0.235 | 0.166 | 0.21 | 0.28 |
| Morbid Obesity |  |  |  |  |  |  |  |  |  |
| HCC 48 | Morbid (Severe) Obesity Due to Excess Calories |  | 0.186 | 0.144 | 0.3 | 0.178 | 0.164 | 0.118 | 0.442 |
| Rheumatoid Arthritis |  |  |  |  |  |  |  |  |  |
| HCC 93 | Rheumatoid Arthritis and Other Specified Inflammatory Rheumatic Disorders |  | 0.617 | 0.47 | 0.439 | 0.384 | 0.405 | 0.288 | 0.297 |
| Dementia |  |  |  |  |  |  |  |  |  |
| HCC 125 | Dementia, Severe |  | 0.341 | 0.296 | 0.438 | 0.367 | 0.401 | 0.345 | 0 |
| HCC 126 | Dementia, Moderate |  | 0.341 | 0.296 | 0.438 | 0.367 | 0.401 | 0.345 | 0 |
| HCC 127 | Dementia, Mild or Unspecified |  | 0.341 | 0.296 | 0.438 | 0.367 | 0.401 | 0.345 | 0 |
| Heart Disease |  |  |  |  |  |  |  |  |  |
| HCC 222 | End Stage Heart Failure |  | 2.505 | 5.77 | 2.927 | 6.612 | 3.009 | 6.106 | 0.826 |
| HCC 223 | Heart Failure With Heart Assist Device/Artificial Heart |  | 2.505 | 5.77 | 2.927 | 6.612 | 3.009 | 6.106 | 0.826 |
| HCC 224 | Acute on Chronic Heart Failure |  | 0.36 | 0.442 | 0.406 | 0.537 | 0.311 | 0.411 | 0.217 |
| HCC 225 | Acute Heart Failure |  | 0.36 | 0.442 | 0.406 | 0.537 | 0.311 | 0.411 | 0.217 |
| HCC 226 | Heart Failure, Except End-Stage and Acute |  | 0.36 | 0.442 | 0.406 | 0.537 | 0.311 | 0.411 | 0.217 |
| Stroke |  |  |  |  |  |  |  |  |  |
| HCC 248 | Intracranial Hemorrhage |  | 0.239 | 0.18 | 0.377 | 0.332 | 0.313 | 0.183 | 0.081 |
| HCC249 | Ischemic or unspecified stroke |  | 0.239 | 0.18 | 0.377 | 0.277 | 0.299 | 0.172 | 0.081 |
| Lung Disorders |  |  |  |  |  |  |  |  |  |
| HCC 279 | Severe Persistent Asthma |  | 0.818 | 0.842 | 0.594 | 0.808 | 0.65 | 0.804 | 0.873 |
| HCC 280 | Chronic Obstructive Pulmonary Disease, Interstitial Lung Disorders, and Other Chronic Lung Disorders |  | 0.319 | 0.209 | 0.39 | 0.281 | 0.321 | 0.234 | 0.312 |
| Chronic Kidney Disease |  |  |  |  |  |  |  |  |  |
| HCC 326 | Chronic Kidney Disease, Stage 5 |  | 0.815 | 0.927 | 0.985 | 0.946 | 0.965 | 1.05 | 0.958 |
| HCC 327 | Chronic Kidney Disease, Severe (Stage 4) |  | 0.514 | 0.523 | 0.565 | 0.661 | 0.484 | 0.447 | 0.462 |
| HCC 328 | Chronic Kidney Disease, Moderate (Stage 3B) |  | 0.127 | 0.179 | 0.116 | 0.181 | 0.14 | 0.178 | 0.145 |
| HCC 329 | Chronic Kidney Disease, Moderate (Stage 3, Except 3B) |  | 0.127 | 0.179 | 0.116 | 0.181 | 0.14 | 0.178 | 0.145 |
| Disease Interactions |  |  |  |  |  |  |  |  |  |
| N/A | Diabetes*Heart Failure |  | 0.112 | 0.023 | 0.183 | 0.041 | 0.164 | 0.053 | 0.209 |
| N/A | Heart Failure*Chronic Lung Disorder |  | 0.078 | 0.062 | 0.109 | 0.097 | 0.14 | 0.108 | 0.145 |
| N/A | Heart Failure*Kidney |  | 0.176 | 0.314 | 0.194 | 0.42 | 0.14 | 0.328 | 0 |
| N/A | Chronic Lung Disorder*Cardiorespiratory Failure |  | 0.254 | 0.242 | 0.34 | 0.275 | 0.329 | 0.27 | 0.331 |

**Notes:**
CNA: Community Non-dual aged, CND: Community Non-dual disabled, CFA: Community Full Benefit dual aged, CFD: Community Full Benefit dual disabled, CPA: Community Partial Benefit dual aged, CPD: Community Partial Benefit dual disabled, INS: Institutionalized.

Notably, diabetes is one of two disease groups (the other being congestive heart failure) for which the coefficients are “constrained” in the HCC V28 model, meaning that the same coefficient is applied to three different HCCs: HCC 36, “diabetes with severe acute complications”; HCC 37, “diabetes with chronic complications”; and HCC 38, “diabetes with glycemic, unspecified, or no complications.” Thus, by using the V28 model, diabetes presence is particularly straightforward in that it could be based on a clinically objective measure (e.g., HbA1C) and would not be subject to additional clinical discretion regarding severity of complications. However, the constrained coefficient may not adequately address the full range of cost savings that could vary across these diabetes severity subgroups, or across subgroups eligible for AOMs under this model. Additional analysis could verify the applicability of the HCC coefficients to the population targeted by this intervention and, if appropriate, CMS could explore using differentiated coefficients across these three diabetes severity categories, as was done in previous versions of the HCC model.

CMS also has a risk-adjustment model based on hierarchical codes for prescription drug costs— CMS-RxHCC—that adjusts for many of the same conditions as the Part C model, with some differences. For example, CMS-RxHCC adjusts for hypertension whereas the Part C model does not. The implication is that a diagnosis of hypertension will impact prescription drug costs, but it won’t have a significant impact on overall non-drug medical costs. Conversely, the Part C model adjusts for stroke whereas CMS-RxHCC does not. While below we provide estimates of savings associated only with the Part C model, a VBA should also include savings associated with Part D, which could be measured in a similar way.

### Avoided Cases

The next component of the risk-based payment is the number of cases avoided for the relevant conditions, *c*. To keep things transparent, we propose computation using historical controls.^2^ Assume we have *τ* months of historical data before the adoption of the GLP-1 contract. Define *α*_*ctda*_ as the actual historical prevalence rate, per 1,000 for condition *c*, demographic group *d*, age group *a*, at time *t*. Assuming the historical data begins in historical month *H*and ends in month *H* + (*τ* − 1), then define the historical baseline incidence rates:

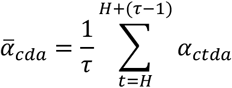

Assume the contract begins in month *H*and ends in month *H*+ *T*. The actual monthly incidence rate, *α*_*ctda*_, during the contract period *t* ∈ {*H*, …, *H*+ *T*} can be observed. Now define 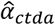 as the number of avoided cases of condition *c*, at time *t*, for patients of age *a* in demographic group *d*, where

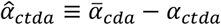

Notice that avoided cases can be negative if GLP-1 treatment is associated with more cases than in the historical controls.

### Contracted Payment Amount

In each period *t*, the manufacturer receives (or pays if the amount is negative) the amount:

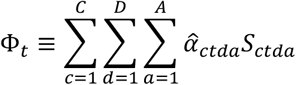

Consider a contract that covers GLP-1 drugs for all clinically eligible Medicare beneficiaries such that manufacturers will receive 50% of the resulting savings from diabetes, heart disease, stroke and cancer prevention.^3^ To provide some context, we report the projected cases avoided based on our prior microsimulation analysis of the effects of broader GLP-1 usage in the Medicare population. Specifically, the prior work studies a counterfactual scenario where Medicare (but not private insurers) expands access to all clinically eligible patients (BMI ≥ 30 kg/m^2^, or BMI ≥ 27 kg/m^2^ in the presence of at least one weight-related comorbidity) compared to the status quo of no Medicare coverage of GLP-1s for weight loss.[8] Table 3 shows the “status quo” and “expanded access” disease prevalence rates for diabetes and morbid obesity in traditional Medicare after 10 years, along with the associated total number of cases. Mediation analysis of clinical trial results for tirzepatide shows that 39% to 55% of the reduction in the risk of diabetes was mediated through the reduction in body weight [7]. This implies that our prior microsimulation results, which estimate the reduction in diabetes from weight loss alone, would underestimate diabetes cases avoided and the resulting outcomes-based payments from Medicare. We account for the full impact of GLP-1s in diabetes prevention by using the clinical trial results.

**Table 3.** Calculating Manufacturer Bonus Payments From Medicare TM VBA.

|  | Prevalence |  | Cases |  |  | VBA Savings |  |
| --- | --- | --- | --- | --- | --- | --- | --- |
|  | Status Quo | Expanded Access | Status Quo | Expanded Access | Avoided | HCC Coefficient | Monthly Medicare Savings |
| Morbid Obesity (BMI 40+) | 0.08 | 0.02 | 2,679,720 | 777,119 | 1,902,601 | 0.205 | \$ 435,302,445 |
| Diabetes* | 0.28 | 0.03 | 9,379,020 | 937,902 | 8,441,118 | 0.169 | \$ 1,592,936,884 |
| Monthly Medicare Savings | | | | | | | \$ 2,028,239,329 |
| Annual Medicare Savings | | | | | | | \$ 24,338,871,945 |
| Annual Payment to Manufacturers ($\sigma = 50\%$) | | | | | | | \$ 12,169,435,973 |

**Notes:**
\* Uses the diabetes prevention results from clinical trials to capture the full effect of GLP-1s.
Assumes 68.5 million Medicare beneficiaries of which 48.9% are enrolled in TM.
Our prior analysis did not estimate the reduction in disease prevalence by beneficiary eligibility criteria, benefit type or nursing home status. Thus, we use a weighted average of the community, non dual, aged (CAN) coefficient and the community, dual full benefit, aged coefficient (CFA) assuming 17% of beneficiaries are dual eligible.
We assume average monthly TM spending of \$1,114.

To estimate the savings associated with reductions in diabetes and morbid obesity, we used the CMS-HCC model coefficients for Part C, multiplying these by the total number of cases avoided. However, our prior results are not stratified by eligibility status, benefit type and nursing home status. Therefore, we take a simple, conservative approach to reconciling these differences by assuming these disease-prevalence reductions would occur exclusively among non-institutionalized beneficiaries (i.e., using smaller HCC coefficients) and that 17% of the beneficiaries, and thus the disease-incidence reductions, are dual eligible. The coefficients listed in table 3 are a weighted average of the community, non-dual, aged coefficient and the community, dual full benefit, aged coefficient. As described above, the estimates in table 3 do not include related Part D savings.

Using the current traditional Medicare population of 33.5 million beneficiaries, table 3 calculates the monthly savings associated with the reduction in each health condition and thus the total savings that would accrue to GLP-1 manufacturers under a VBA in the 10th year of GLP-1 access.

Under the hypothetical contract, traditional Medicare spending would be reduced by $2 billion a month or $24.3 billion in year 10, and manufacturers would receive $12.2 billion in bonus payments. Reductions in diabetes provide the largest savings.

While not included in the discussion above, Medicare Advantage plans could also engage in value-based contracting for GLP-1s for weight loss such that all Medicare beneficiaries received coverage. If Medicare Advantage enrollees saw similar reductions in the obesity-related conditions included in table 3, we estimate that total Medicare spending in year 10 could fall by as much as $15.5 billion.

Although the example using our prior analysis is useful in demonstrating how Medicare could structure a VBA contract, it has several limitations. First, the impact that GLP-1s have on disease prevalence will grow over time, such that savings in the first few years of the hypothetical contract are likely to be much smaller. Additionally, the disease-reduction estimates assume that 100% of clinically eligible patients use GLP-1s and remain adherent to the drugs for 10 years. Real-world experiences with GLP-1s may be different, which would result in potentially smaller reductions in disease prevalence. However, this would also be associated with lower payments owed to manufacturers. CMS coverage may also limit coverage to patients with a BMI > 30, whereas our prior Medicare analysis was consistent with the Food and Drug Administration obesity indication and extended treatment to patients with a BMI ≥ 27 in the presence of at least one weight-related comorbidity (hypertension or type 2 diabetes). Treating a smaller patient population may impact the avoided cases of each disease but would also reduce associated costs of treatment. Finally, obesity impacts more health conditions than we previously modeled, so future analysis should build out the contract to consider new diseases— such as dementia and rheumatoid arthritis—and cost savings to Medicare Part D through the CMS-RxHCC model that are not included in table 3.

## Conclusion

Expanding coverage of GLP-1s for weight loss could help curb the long-term social and economic burden of obesity, which affects millions of Americans and drives rising healthcare costs. However, uncertainty around how the drugs will perform in real-world populations and over the long term has made many insurers reluctant to expand coverage. Alternative payment structures like VBAs offer a solution to insurer concerns by allowing the risks of performance to be shared with manufacturers.

We propose a value-based demonstration in traditional Medicare that would provide access to GLP-1s for patients with weight loss. We outline a straightforward, easy-to-implement contract where CMS can negotiate a lower upfront payment for GLP-1s by sharing a portion of the future Medicare savings. This approach relies on existing risk-adjustment models for Medicare Parts C and D to estimate the savings for obesity-related conditions. Although key contract parameters will need to be negotiated, this structure provides a value-based, feasible method to obtain trillions in health benefits [9] in a fiscally responsible manner.

## Data Availability

A technical appendix providing a detailed description of the microsimulation model and all data sets is available upon request.

## Footnotes

1 In the lexicon of former U.S. Defense Secretary Donald Rumsfeld, these are “known unknowns.”

2 In the long run, it will become impractical to rely on “pre-VBA” historical controls to compute cases avoided. Two possible strategies for long-run contracting include: traditional volume-based contracting, relying on the accumulation of real-world data on the actual health benefits of GLP-1 drugs for weight loss; or VBAs in which manufacturers pay for incident cases rather than being paid for cases avoided. The latter approach works better in the presence of more accurate long-run estimates of “expected value,” which could serve to anchor a base payment from which “disease incidence penalties” would be deducted.

3 Research suggests that obesity impacts additional health conditions, but our prior research on the benefits of Medicare coverage for AOMs estimates the decline in prevalence of these specific conditions as the result of expanded coverage.

